# Arterial Stiffness is an Important Predictor of Heart Failure with Preserved Ejection Fraction - The Effects of Phosphate Retention-

**DOI:** 10.1101/2023.08.14.23294103

**Authors:** Yuji Mizuno, Toshifumi Ishida, Kenichi Tsujita, Michihiro Yoshimura

## Abstract

**BACKGROUND:** Heart failure with preserved ejection fraction (HFpEF) is a major health concern. There is a growing recognition of the causal interplay between arterial stiffness and HFpEF. We recently reported that the deterioration of phosphate homeostasis is a trigger for both arterial stiffness. This study focuses on whether arterial stiffness due to phosphate retention could be a predictor for HFpEF.

**METHODS:** The study subjects were 158 patients (68 males and 90 females, mean age 74.8±11.2). They received echocardiography, central blood pressure (BP) and blood biochemistry tests. HFpEF was defined according to the guidelines of the European Society of Cardiology 2021. Pulse wave velocity (PWV) and central systolic blood pressure (CSBP) were used as markers for arterial stiffness and cardiac afterload, respectively. We measured serum levels of fibroblast growth factor 23 (FGF23) as markers of phosphate retention.

**RESULTS:** The serum levels of FGF23 had significant relationship with PWV (t=3.33, p<0.001). PWV had a significant positive relationship with CSBP (t=4.54, p<0.001). PWV furthermore had significant relationships with LV mass index (t=4.74), plasma BNP levels (t=5.44), and relative wall thickness (t=3.83), e’ (t=-4.21) and E/e’ (t=7.88) (p<0.001, respectively). Multivariate logistic regression analysis using independent factors, including PWV higher values, sex and hypertension, revealed that PWV higher values (t=5.89, p<0.0001) and hypertension (t=2.17, p=0.031) were significant predictors for the dependent factor (HFpEF).

**CONCLUSIONS:** Arterial stiffness amplified cardiac afterload, leading to LV concentric hypertrophy and LV diastolic dysfunction. This study presents that arterial stiffness due to phosphate retention, and hypertension are important predictors of HFpEF.

**What is New?:** Arterial stiffness is an important predictor for HFpEF. Arterial stiffness is caused by an increase in phosphate retention due to aging and CKD via kidney nephron loss. Arterial stiffness amplifies cardiac afterload leading to LV concentric hypertrophy and LV diastolic dysfunction.

**What are the Clinical Implications?:** Arterial calcification and/or stiffness should be a new target in cardiovascular diseases. Phosphate is thought to be an important aging accelerating factor. Phosphate regulating medications, phosphate restriction diets, and osteoporosis therapies may all be protective against aging related diseases including HFpEF.

---

Heart failure with preserved ejection fraction (HFpEF) is a world-wide health concern. The number of patients with HFpEF is increasing as populations age. The mechanism though is unclear.^1-4^ There is a growing recognition of the causal interplay between arterial stiffness and diastolic dysfunction and/or HFpEF.^5-14^ It has not been widely known what the main cause of arterial stiffness is, nor how it leads to the pathological mechanism of HFpEF.

Hemodialysis patients present the most representative model of HFpEF,^9-11^ as they present the following characteristics: renal dysfunction; arterial calcification; left ventricular (LV) concentric hypertrophy; LV diastolic dysfunction; hypertension; osteoporosis; and. accelerated aging.^9-11,15^

The Klotho/Fibroblast growth factor 23 (FGF23) axis has been focused on as an important regulating factor of accelerated aging, including cardiovascular mortality.^10,16-20^ In response to phosphate intake, FGF23 is secreted mainly from the bones, circulates in the blood, and binds to the Klotho-FGF receptor complexes expressed in the kidneys to promote urinary phosphate excretion.^10,17-20^ The FGF23-Klotho endocrine system is indispensable for maintaining phosphate homeostasis. Loss-of-function mutations in either FGF23 or Klotho cause phosphate retention phenotypes including ectopic calcification and hyperphosphatemia.^10,17-20^ Serum phosphate level is controlled within a small range, whereas phosphate homeostasis is maintained by a counterbalance between the absorption of dietary phosphate from the intestines and the excretion of phosphate from the blood via the kidneys into the urine.^10,17-20^ Parathyroid hormone (PTH) is also a phosphaturic hormone.^17,19,20^ Like FGF23, PTH exerts phosphaturic activity by suppressing phosphate reabsorption at the renal tubules.^20-23^ In the parathyroid organ, FGF23 suppresses production and secretion of PTH, whereas PTH reciprocally induces FGF23 expression.^20-23^

VD3 is a physiologically important active hormone against atherosclerosis, endothelial dysfunction, inflammation, osteoporosis and LV hypertrophy.^24,25^ VD3 is activated by the kidneys and acts on the intestines to increase the absorption of phosphate and calcium, thereby inducing a positive phosphate balance.^20-25^ VD3 is, however, inhibited by FGF23 in a state wherein the retention of phosphate is in balance.^20-23^ It is therefore commonly agreed that FGF23 is the most potent hormone regulating phosphate homeostasis and phosphate retention.^17,20-23^ Phosphate retention and phosphate regulating hormones could therefore be responsible for pathological arterial calcification.^17,20-23^

We recently reported that the deterioration of phosphate homeostasis is an important trigger for arterial stiffness, and that arterial stiffness intensifies cardiac afterload.^26^ It has not yet been reported whether arterial stiffness could lead to HFpEF. To understand the pathological mechanism of HFpEF, we investigated whether arterial stiffness could be a predictor for HFpEF.

## METHODS

### Study Subjects

This is a prospective cross-sectional study comprised of 158 consecutive Japanese patients with Table 1, New York Heart Association Classification II-III, 68 males and 90 females, with a mean age of 74.8 (± 11.2) who were admitted at, or referred to, our institution or out-patient clinic between May, 2018 and March, 2022. They presented, or had presented with, the following: 1) Symptoms (dyspnea, fatigue and fluid retention) and signs of heart failure; 2) Left ventricular ejection fraction (LVEF) ≥50%; and, 3) Objective evidence of cardiac structural and/or functional abnormalities consistent with the presence of LV diastolic dysfunction such as raised LV filling pressures, including early diastolic mitral flow velocity (E)/tissue annular motion velocity (e ’) or E/e ’≥15 on echocardiogram and elevated plasma BNP levels ≥35 pg/mL in sinus rhythm (SR) and ≥105 pg/mL in atrial fibrillation (AF) according to the guidelines of 2021 ESC.^4^ Patients with acute decompensated heart failure (LVEF < 50 %), acute myocardial infarction, acute inflammatory disease, pericardial disease, severe valvular heart disease, congenital heart disease, and infiltrative cardiomyopathies were excluded from the study. This study was conducted in accordance with the Declaration of Helsinki and approved by the ethical committee of our institution.

**Table 1.**
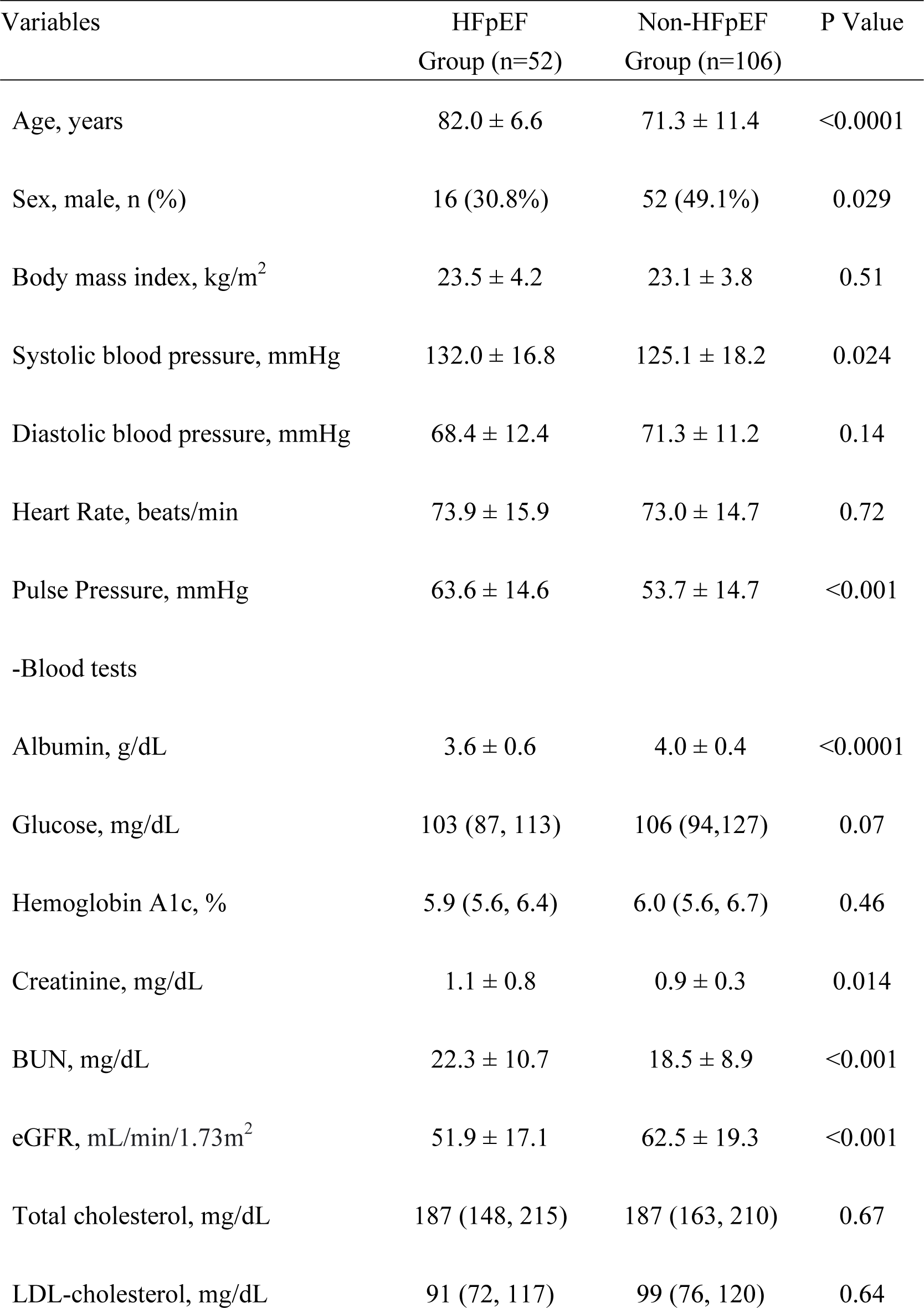
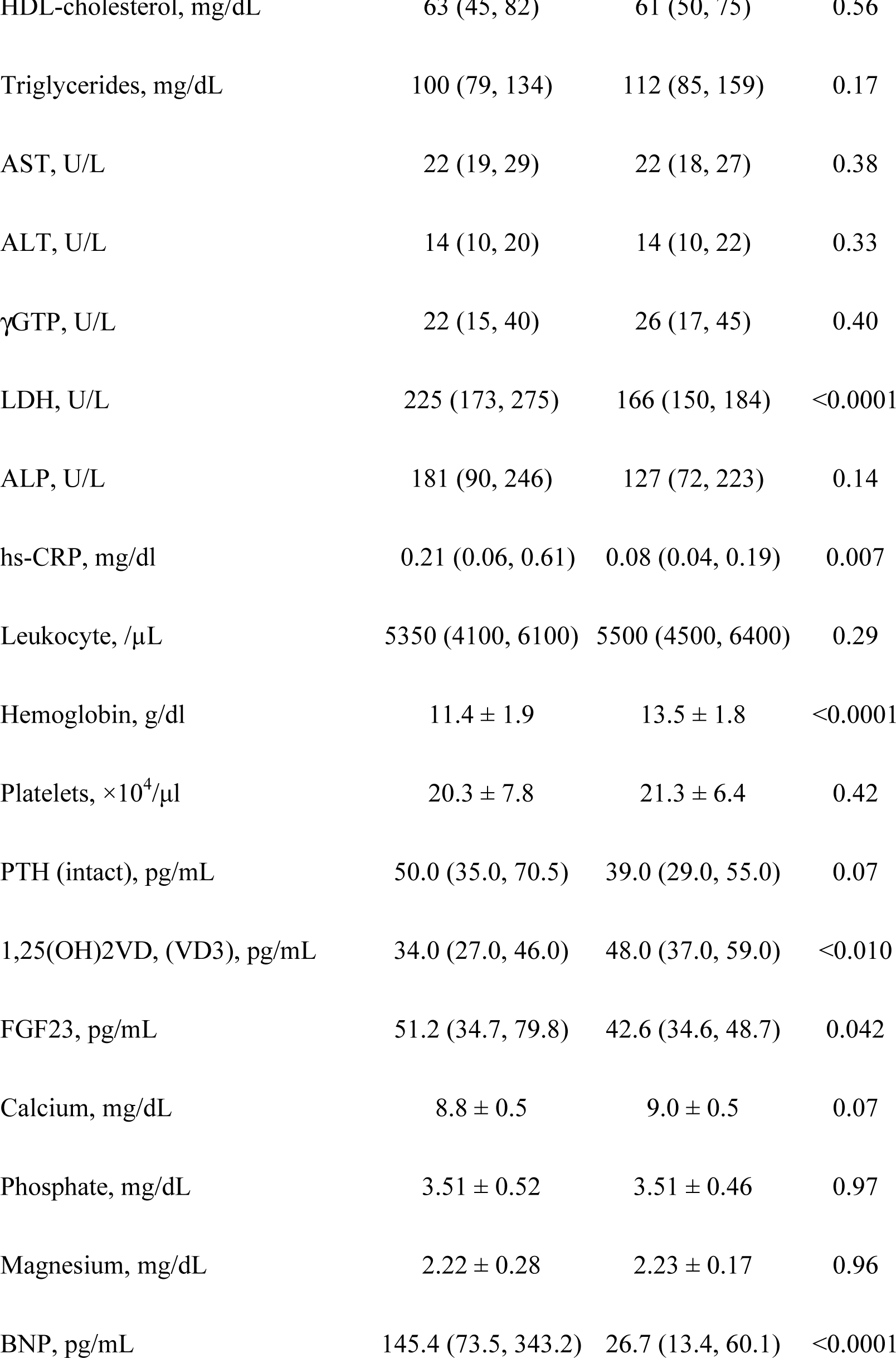
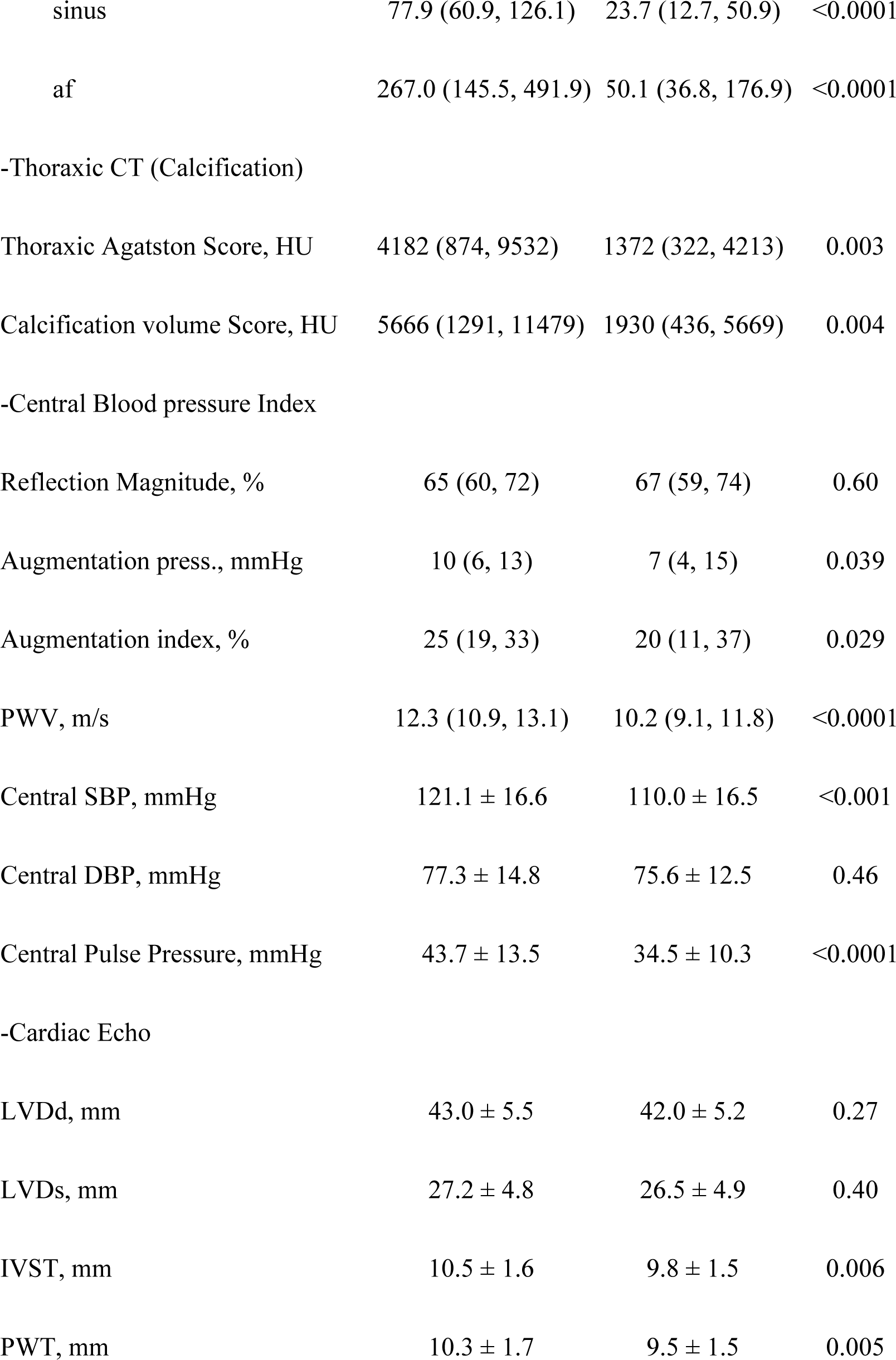
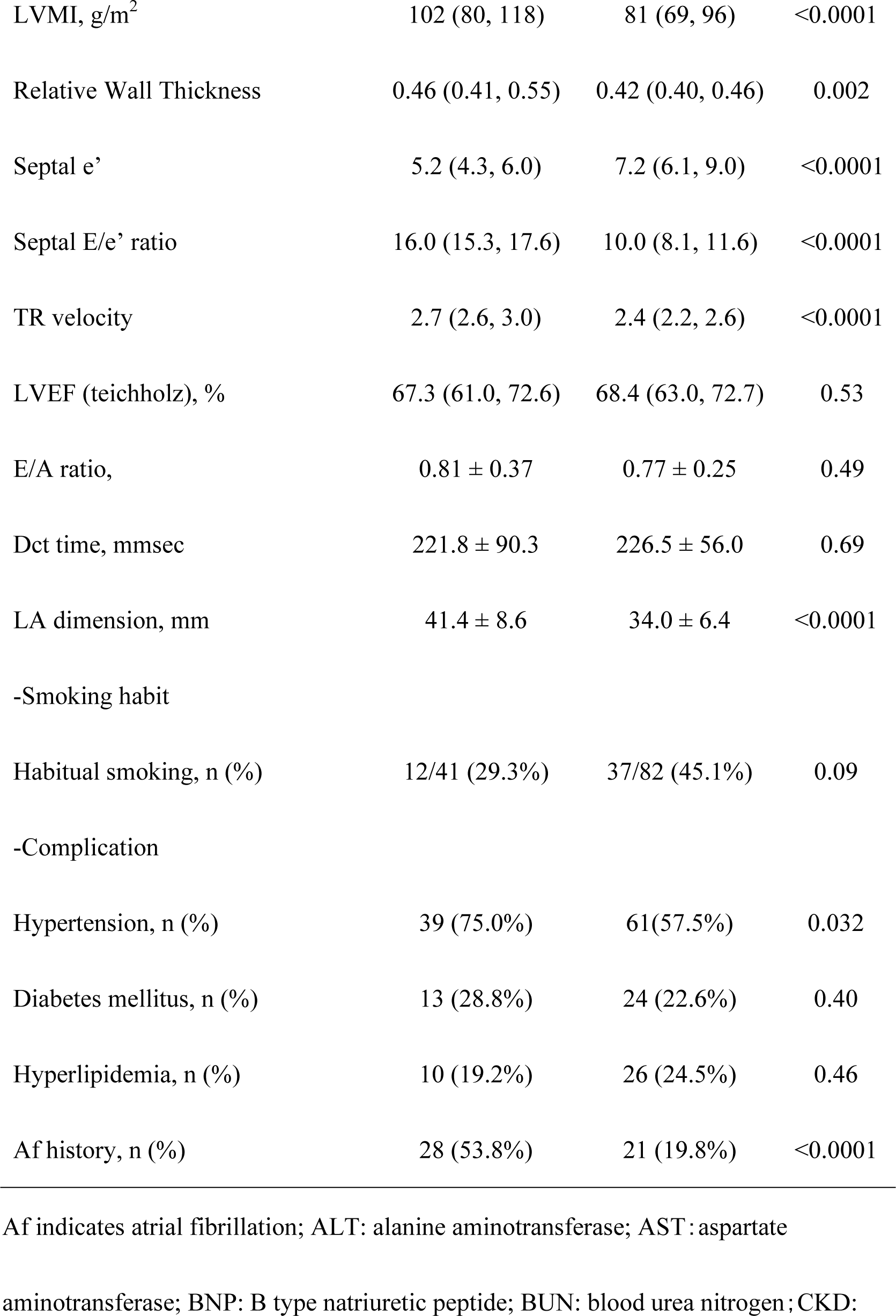
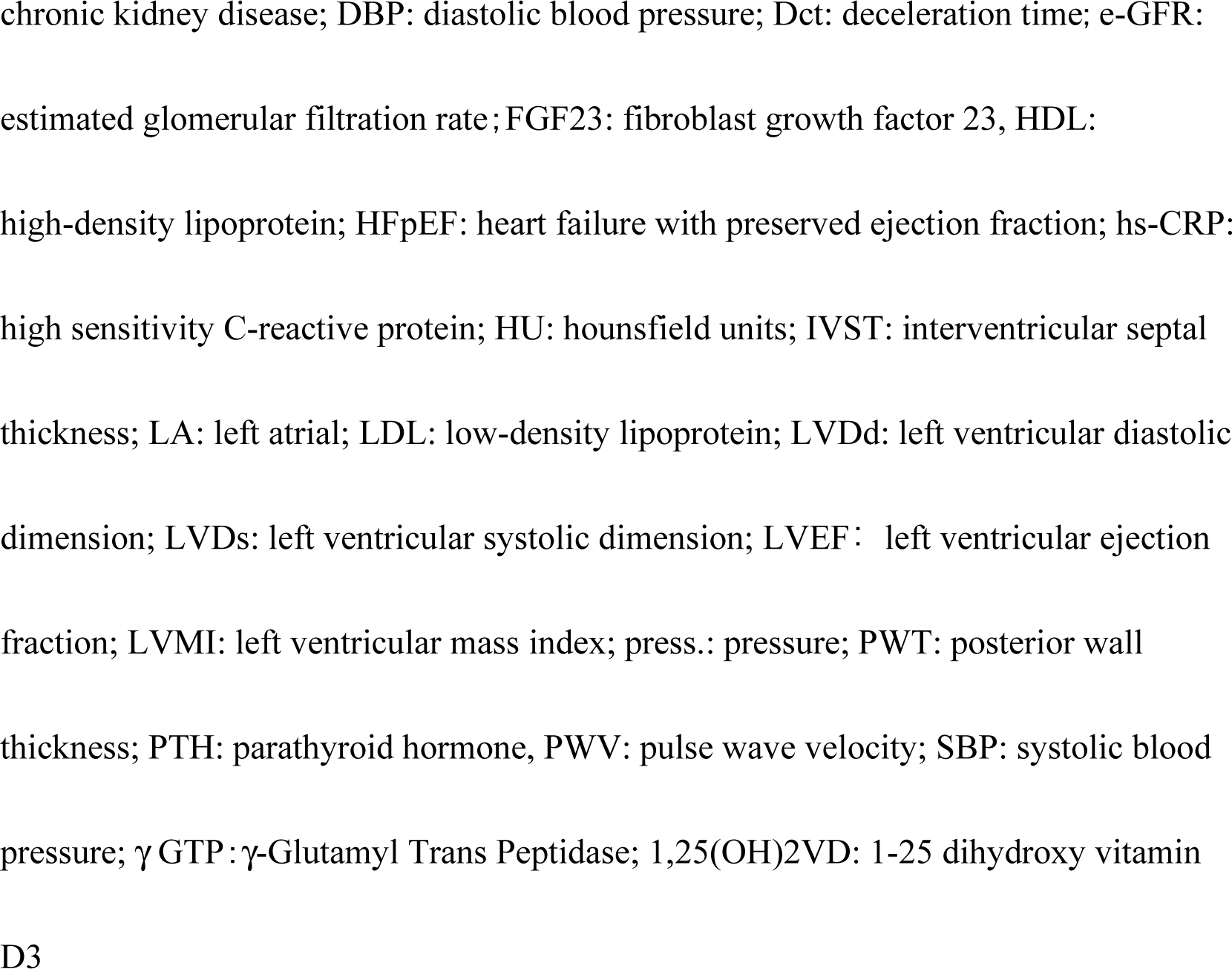
Comparison between HFpEF group and Non-HFpEF group.

### Echocardiography

Echocardiography including two-dimensional, pulse and continuous wave Doppler, color flow Doppler, and tissue Doppler imaging were performed using the iE33 Ultrasound System (Philips Ultrasound Co. Bothell, U.S.A.) with the patients stable at the time of examination either as an ambulatory outpatient or inpatient on the same day or within 2 days of blood sampling for BNP levels. LV and atrial linear dimensions were measured from two-dimensional echocardiographic images and peak E-wave and A-wave velocity, E/A, E/e’, LV diastolic dimension (LVDd), LV systolic dimension (LVDs), LV mass index (LVMI), left atrial dimension (LAD), right ventricular dimension (RVD), pulmonary artery systolic pressure (PASP), interventricular-septal thickness (IVST), posterior-wall thickness (PWT), relative wall thickness (RWT), stroke volume (SV), and LVEF were measured and calculated according to the recommendations of the American Society of Echocardiography and the European Association of Echocardiography.^27^ LV mass was estimated using linear measurements from two dimensional images and indexed to body surface area as LVMI. LV geometry was classified based on RWT, defined as (2×diastolic posterior wall thickness)/LVDd) and LVMI: normal-RWT ≤0.42 and no LV hypertrophy (LVH); eccentric hypertrophy-RWT ≤0.42 and LVH; concentric remodeling-RWT >0.42 and no LVH; concentric hypertrophy-RWT >0.42 and LVH.^27^ Echocardiography was performed by experienced sonographers who were unaware of the clinical information on each patient.

### Assessments of Pulse Wave Velocity and Central Blood Pressure Measurements

This study used pulse wave velocity (PWV) as a marker of arterial stiffness.^14,28^ PWV and central blood pressure related parameters were detected through the Mobil-O-Graph pulse wave analysis (PWA)/ambulatory BP monitoring device (I.E.M. GmBH, Stolberg, Germany) which performs a cuff-based oscillometric method of measurement. The device was approved for blood pressure measurement by the British Hypertension Society and the European Society of Hypertension, and device reliability was demonstrated in comparisons through invasive and non-invasive methods for PWA.^29,30^ Blood pressure (BP) measurements were done on patients’ left upper arms in a sitting position after a 10-minute rest, with the patients left elbows flexed and supported at the heart level on the chair. Augmentation pressure, augmentation index (AI), and central BP including general brachial artery BP measurements were measured.^29,30^ Blood pressure: brachial BP was measured with an Omuron HEM 705-CP semiautomatic oscillometric recorder, using the mean of 3 BP values in the echocardiographic laboratory. Pulse pressure (PP) was calculated as systolic BP minus diastolic BP.

### Thoracic Aortic Calcification (TAC) Scores

TAC burden was measured from each participant’s computed tomographic scan, and TAC was from the aortic annulus, above the aortic valve, to the lower edge of the pulmonary artery bifurcation (ascending aorta), and from the lower edge of the pulmonary bifurcation to the cardiac apex (descending aorta). The TAC score was determined for each study participant using the Agatston score and calcification volume score, which have been widely used in the scientific literature as a convenient TAC quantification method.^31^

### Blood Chemistry Measurements

Blood samples for measurement of clinical chemistry and other data were collected from the patients in a supine position after an overnight fast. The biochemical and other analyses were done using standard laboratory procedures. Venous blood samples were obtained at enrollment, processed, and then stored at -80°C until time of assay.

Serum FGF23 levels were measured by an enzyme-linked immunosorbent assay (ELISA) that recognizes only full-length biologically active FGF23 with a detection limit of 3 pg/ml (Kainos, Japan). The reference range of FGF23 in healthy adults measured by this ELISA is 10-50 pg/ml with a mean value of about 30 pg/ml.^32^

Levels of an active form of Vitamin D, 1,25(OH)2D: (VD3) were determined at baseline with a fully automated and sensitive immunoassay that uses a recombinant fusion construct of the vitamin D receptor ligand binding domain for the specific capture of VD3 (DiaSorin, Saluggia, Italy). The limit of quantification for this VD3 assay is 5 pg/mL and the reference interval determined in healthy volunteers ranged between 25.0 and 86.5 pg/mL with a median of 48.1 pg/mL.^33^

The intact PTH assay was performed using Allegro Intact PTH (I-Nichols, San Juan Capistrano, CA). Normal values range from 10 to 65 pg/mL.^34^

Plasma BNP levels were measured using a specific immunoradiometric assay for human BNP (TOSOH Corp, Tokyo, Japan).^35^ Minimal detectable quantity of human BNP was 2.0 pg/mL. The mean intra-assay and inter-assay coefficients of variation were 2.3% and 3.0%, respectively.

### Statistical Analysis

The baseline clinical data were expressed as the mean ± SD or median (25th, 75th percentile) for continuous variables, and differences within the group were evaluated with the unpaired *t*-test or the Mann-Whitney rank sum test. For discrete variables, the data were expressed as counts and percentages and analyzed with the Chi square test. Classification of habitual smoking included current and past smokers. Linear regression analysis was used to assess the association between each parameter. PWV levels were divided by the medium of each parameter for logistic analysis. Collinearity was estimated in the selection of independent variables for dependent variable in multivariable logistic analyses.^36^ A two-tailed value of *p* < 0.05 was considered to be statistically significant. The analyses were done using the STATA software program (STATA 17.0, STATA Corp., College Station, TX, U.S.A.).

## RESULTS

### Clinical Characteristics

Table 1 compared the clinical characteristics between the with HFpEF group and the non-HFpEF group. The HFpEF group had the following properties: significantly increased age; female rate; systolic BP; PP; plasma level of creatinine; blood urea nitrogen (BUN); lactate dehydrogenase (LDH); hypersensitive C-reactive protein (hs-CRP); FGF23; and BNP. The HFpEF group also had significantly decreased body mass index (BMI), estimated glomerular filtration rate (eGFR), and plasma levels of albumin, hemoglobin and VD3 compared with the non-HFpEF group. The HFpEF group indicated significantly higher aortic calcification scores, Agatston score and calcium volume score, compared with the non-HFpEF group. The HFpEF group moreover presented an increase in arterial stiffness related parameters: AI; PWV; systolic BP; PP; central systolic BP; and central PP. The HFpEF group had furthermore increased LV thickness: IVST, PWT, RWT, and LV diastolic dysfunction markers including E/é, e’, LAD, and significantly they frequently were receiving hypertension and heart failure medications, and they had a history of atrial fibrillation (AF) compared with the non-HFpEF group. There were no differences as regards the characteristics of obesity and/or DM in this study.

### Correlations Between Each Parameter

Age had a significant positive relationship with PWV, and eGFR had a significant negative relationship with PWV (Figure 1). The serum levels of FGF23 had a significant positive relationship with PWV (t=3.33, p=0.001), and that of VD3 had a significant negative relationship with PWV (t=-2.29, p=0.024).

**Figure 1.**
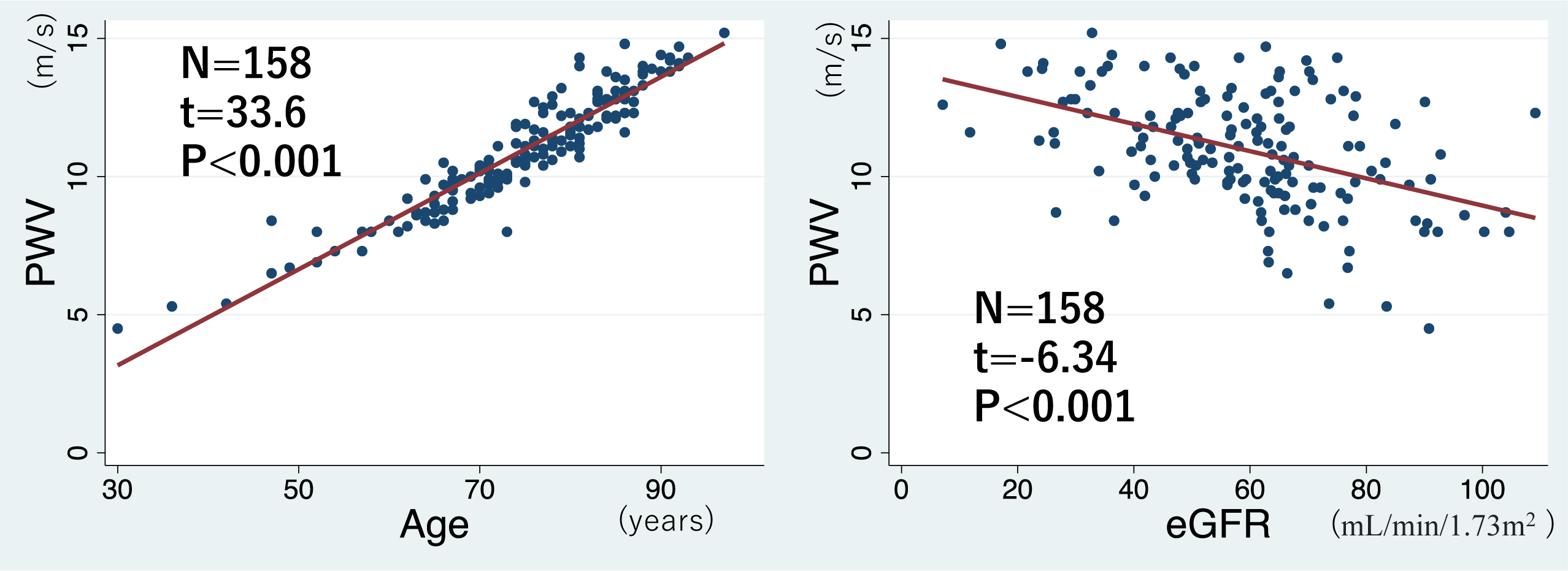
The relationships between PWV and age as well as eGFR. PWV, arterial stiffness marker, shows significant relationships with age and eGFR.

There were significant positive correlations between PWV and augmentation pressure as well as central systolic BP (Figure 2).

**Figure 2.**
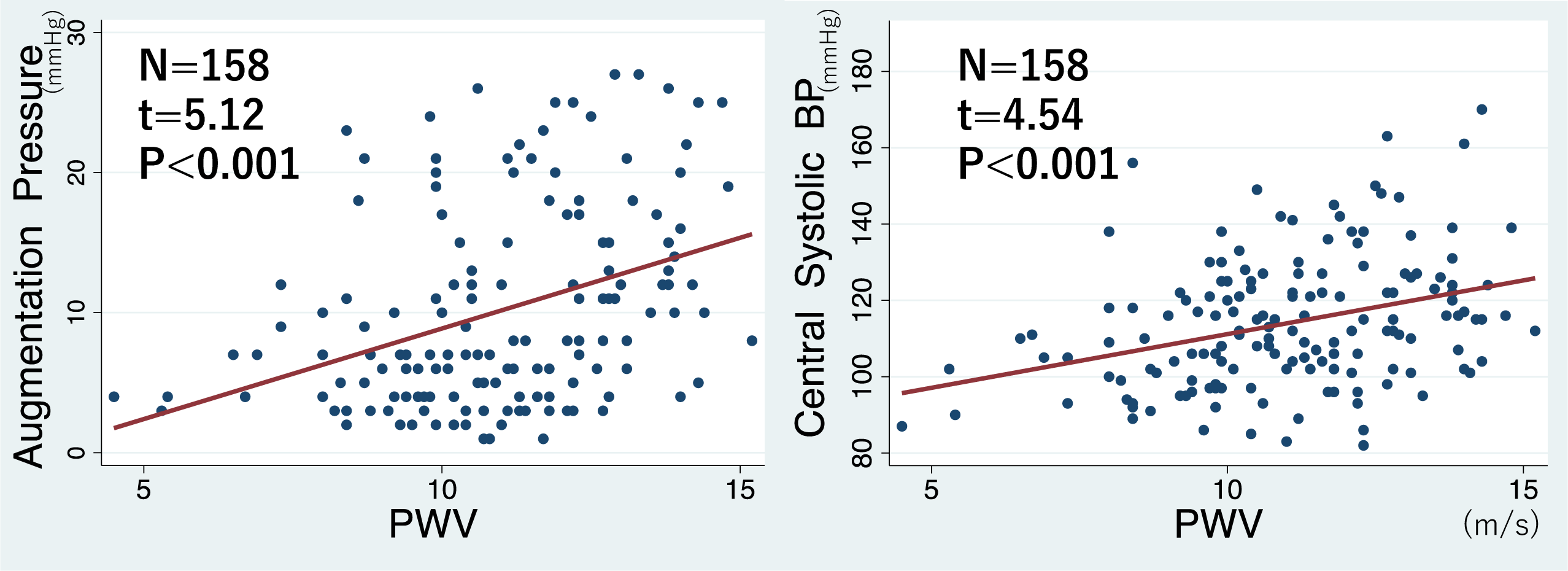
The relationships between PWV and afterload. PWV shows the significant positive relationship with augmentation pressure and central systolic blood pressure that is an indicator of cardiac afterload.

PWV had a significant positive correlation with LV hypertrophic markers: LVMI and plasma levels of BNP. Additionally, it showed a significant positive correlation with RWT, so-called concentric hypertrophy. (Figure 3) PWV finally had significant correlations with e’ and E/e’ ratio. (Figure 4)

**Figure 3.**
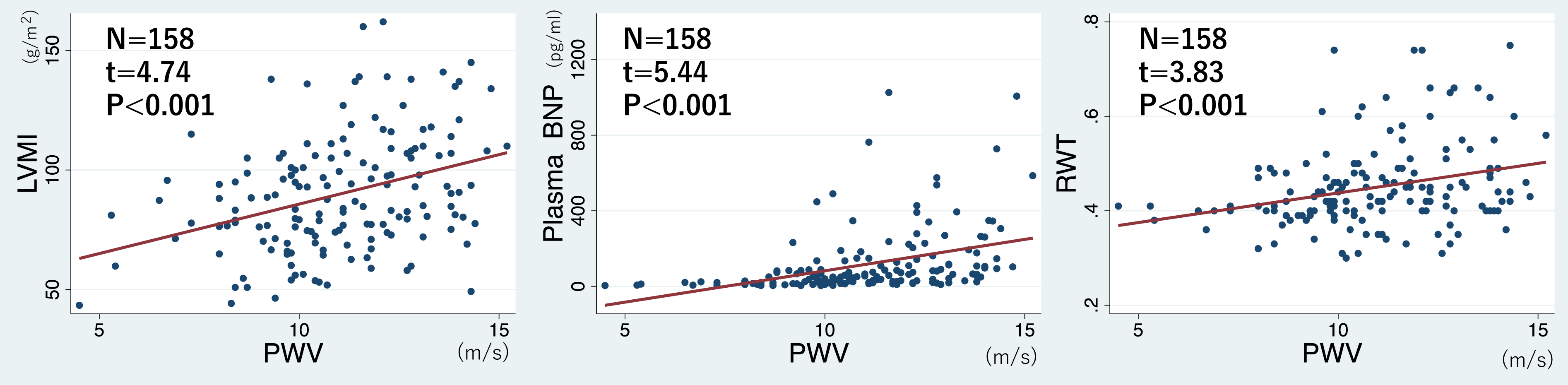
The relationships between PWV and LV hypertrophic markers. PWV showed the significant positive relationship with LV mass index and plasma levels of BNP. PWV had the positive relationship with relative wall thickness, a marker of concentric hypertrophic hypertrophy.

**Figure 4.**
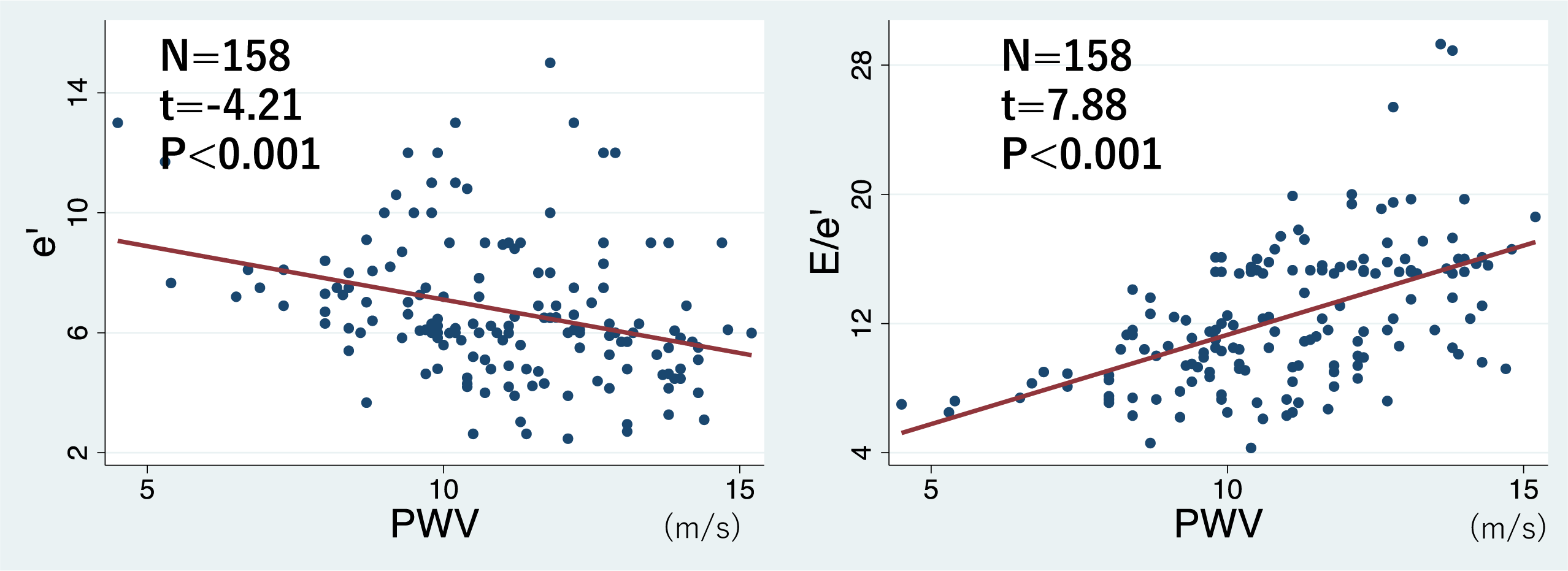
The relationships between PWV and HFpEF judgement parameters. PWV finally had the significant relationships with e’ and E/e’ that are judgement parameters of HFpEF

Central systolic BP had a significant relationship with LVMI (t=3.13, p=0.002), e’ (t=-3.78, p<0.001), and E/e’ (t=2.54, p=0.012).

Females had significantly increased: age (76.7 ± 9.6 vs 72.4 ± 12.6 yrs., p=0.017); serum phosphate level (3.60 ± 0.50 vs 3.36 ± 0.44 mg/dl, p=0.016); central pulse pressure (40.7 ± 12.7 vs 33.5 ± 10.1 mm Hg, p<0.001); AI (25 (17, 38) vs 17 (9, 30) %, p=0.0016); and E/e’ (11.6 (8.5, 15.8) vs 9.0 (6.5, 13.3), p=0.0136). Females had significantly decreased: BMI (22.7 ± 4.4 vs 24.0 ± 3.1 Kg/m^2^, p=0.044); hemoglobin (12.2 ± 1.55 vs 13.6 ± 2.41 g/dl, p<0.001); glucose (99 (91, 113) vs 111 (99, 129) mg/dl, p=0.003); γGTP (20 (14, 28) vs 39 (28, 61) IU/L, p<0.0001); and, hip-bone mineral density (0.69 (0.53, 0.79) vs 0.85 (0.72, 1.01) g/cm^2^, p<0.001). (Table 2)

**Table 2.**
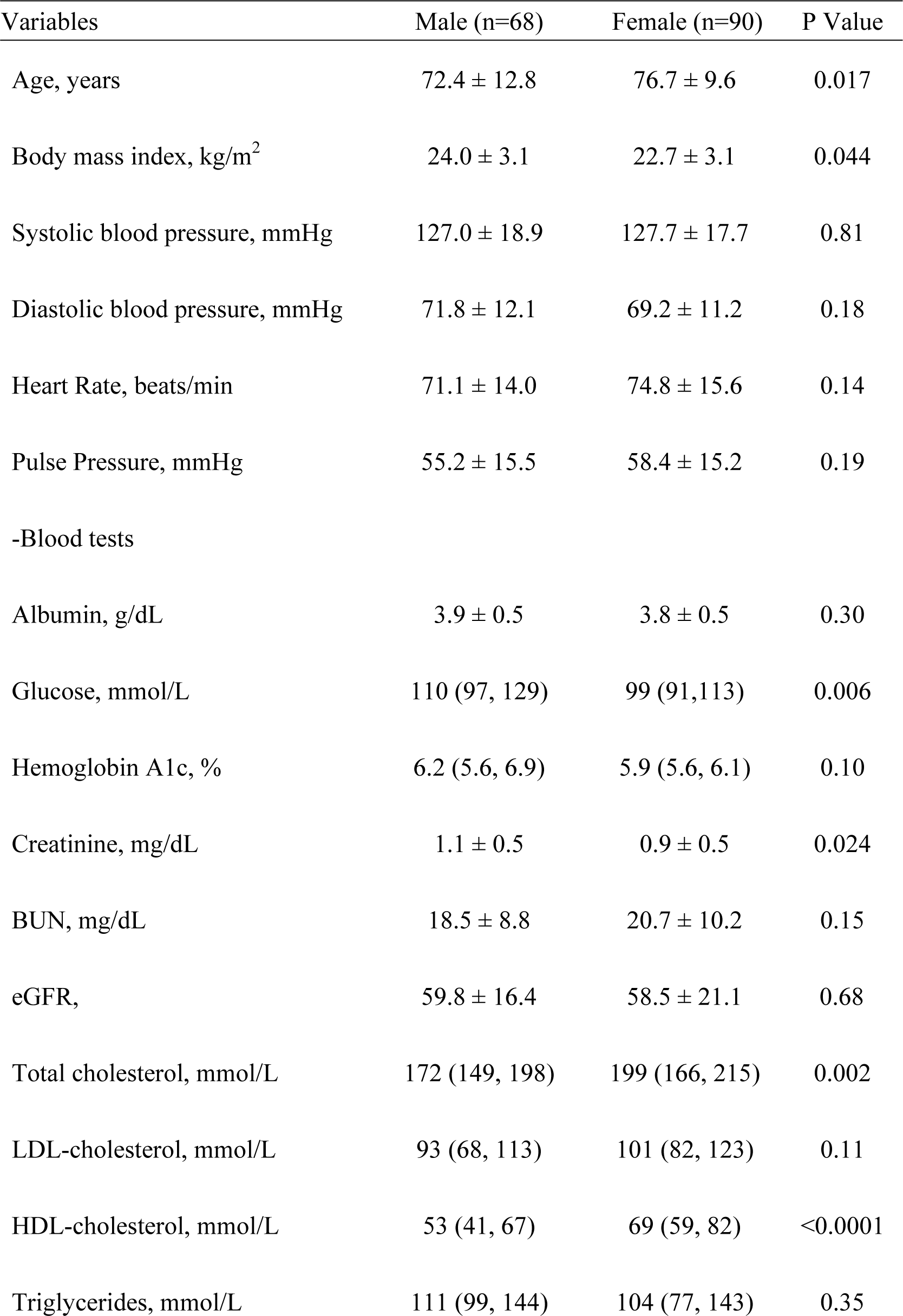
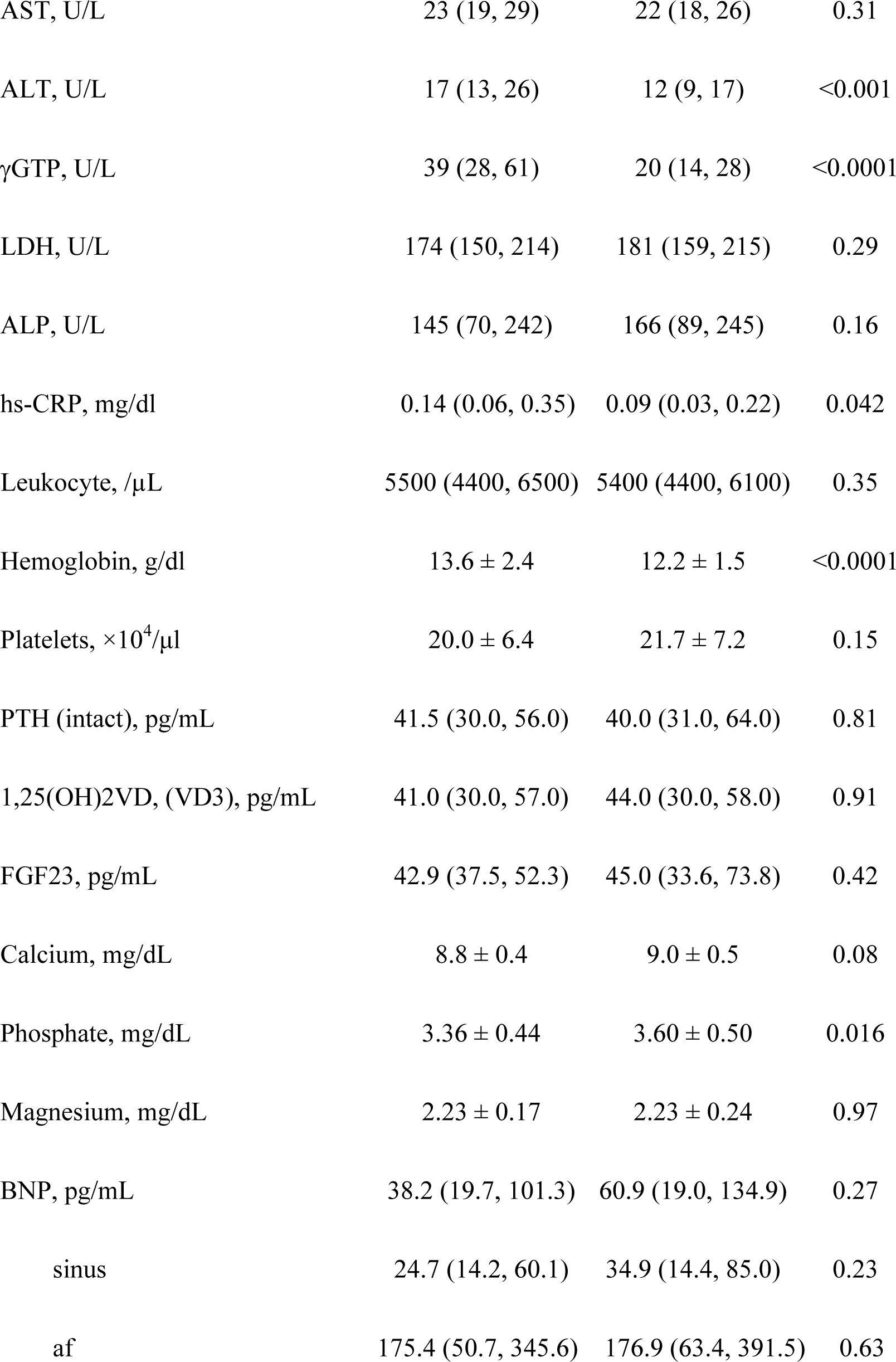
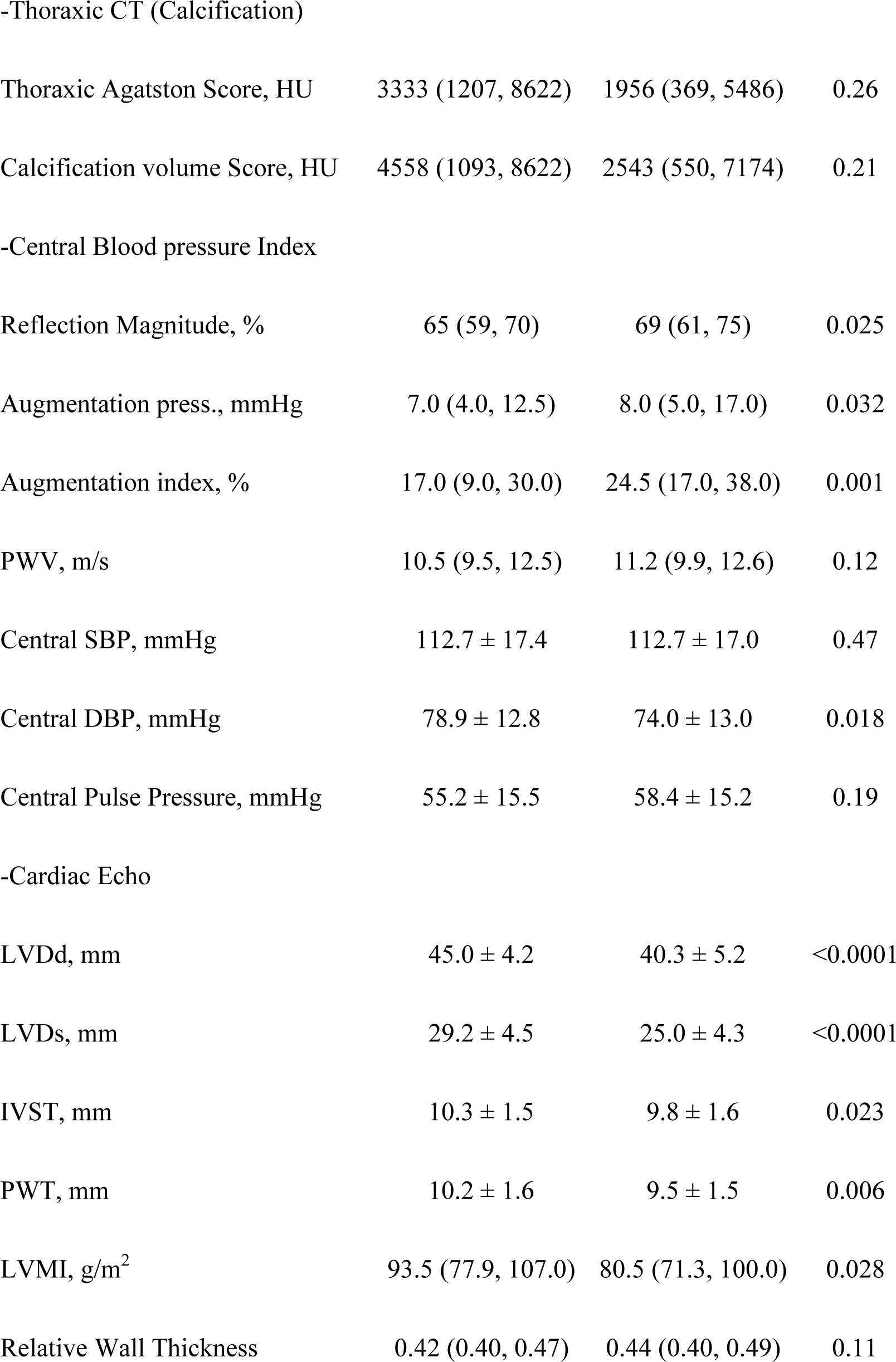
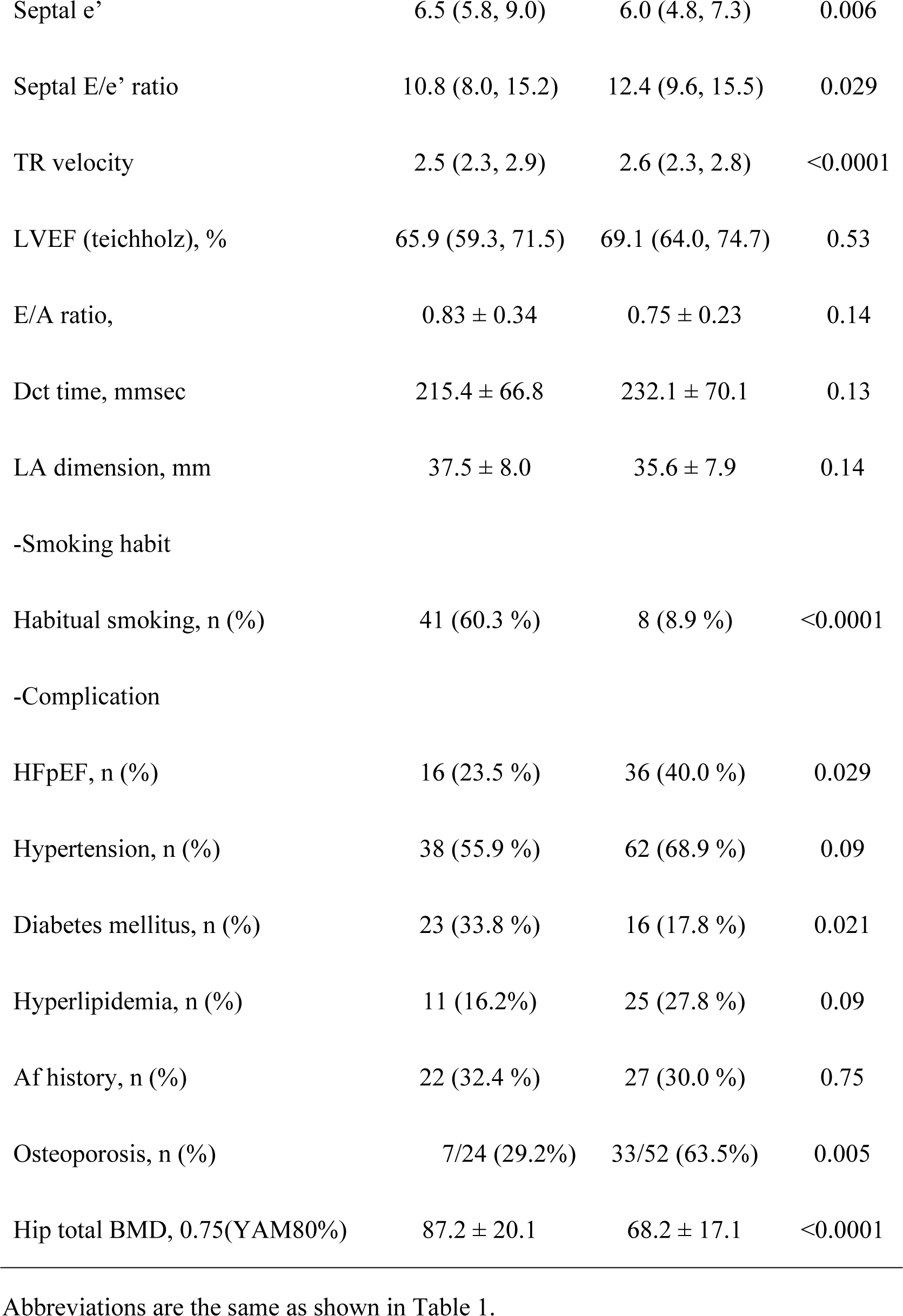
Comparison between Male and Female.

| Variables | Male (n=68) | Female (n=90) | P Value |
| --- | --- | --- | --- |
| Age, years | 72.4 ± 12.8 | 76.7 ± 9.6 | 0.017 |
| Body mass index, kg/m<sup>2</sup> | 24.0 ± 3.1 | 22.7 ± 3.1 | 0.044 |
| Systolic blood pressure, mmHg | 127.0 ± 18.9 | 127.7 ± 17.7 | 0.81 |
| Diastolic blood pressure, mmHg | 71.8 ± 12.1 | 69.2 ± 11.2 | 0.18 |
| Heart Rate, beats/min | 71.1 ± 14.0 | 74.8 ± 15.6 | 0.14 |
| Pulse Pressure, mmHg | 55.2 ± 15.5 | 58.4 ± 15.2 | 0.19 |
| -Blood tests |  |  |  |
| Albumin, g/dL | 3.9 ± 0.5 | 3.8 ± 0.5 | 0.30 |
| Glucose, mmol/L | 110 (97, 129) | 99 (91,113) | 0.006 |
| Hemoglobin A1c, % | 6.2 (5.6, 6.9) | 5.9 (5.6, 6.1) | 0.10 |
| Creatinine, mg/dL | 1.1 ± 0.5 | 0.9 ± 0.5 | 0.024 |
| BUN, mg/dL | 18.5 ± 8.8 | 20.7 ± 10.2 | 0.15 |
| eGFR, | 59.8 ± 16.4 | 58.5 ± 21.1 | 0.68 |
| Total cholesterol, mmol/L | 172 (149, 198) | 199 (166, 215) | 0.002 |
| LDL-cholesterol, mmol/L | 93 (68, 113) | 101 (82, 123) | 0.11 |
| HDL-cholesterol, mmol/L | 53 (41, 67) | 69 (59, 82) | <0.0001 |
| Triglycerides, mmol/L | 111 (99, 144) | 104 (77, 143) | 0.35 |
| AST, U/L | 23 (19, 29) | 22 (18, 26) | 0.31 |
| ALT, U/L | 17 (13, 26) | 12 (9, 17) | <0.001 |
| γGTP, U/L | 39 (28, 61) | 20 (14, 28) | <0.0001 |
| LDH, U/L | 174 (150, 214) | 181 (159, 215) | 0.29 |
| ALP, U/L | 145 (70, 242) | 166 (89, 245) | 0.16 |
| hs-CRP, mg/dl | 0.14 (0.06, 0.35) | 0.09 (0.03, 0.22) | 0.042 |
| Leukocyte, /μL | 5500 (4400, 6500) | 5400 (4400, 6100) | 0.35 |
| Hemoglobin, g/dl | 13.6 ± 2.4 | 12.2 ± 1.5 | <0.0001 |
| Platelets, ×10<sup>4</sup>/μl | 20.0 ± 6.4 | 21.7 ± 7.2 | 0.15 |
| PTH (intact), pg/mL | 41.5 (30.0, 56.0) | 40.0 (31.0, 64.0) | 0.81 |
| 1,25(OH)2VD, (VD3), pg/mL | 41.0 (30.0, 57.0) | 44.0 (30.0, 58.0) | 0.91 |
| FGF23, pg/mL | 42.9 (37.5, 52.3) | 45.0 (33.6, 73.8) | 0.42 |
| Calcium, mg/dL | 8.8 ± 0.4 | 9.0 ± 0.5 | 0.08 |
| Phosphate, mg/dL | 3.36 ± 0.44 | 3.60 ± 0.50 | 0.016 |
| Magnesium, mg/dL | 2.23 ± 0.17 | 2.23 ± 0.24 | 0.97 |
| BNP, pg/mL | 38.2 (19.7, 101.3) | 60.9 (19.0, 134.9) | 0.27 |
| sinus | 24.7 (14.2, 60.1) | 34.9 (14.4, 85.0) | 0.23 |
| af | 175.4 (50.7, 345.6) | 176.9 (63.4, 391.5) | 0.63 |
| -Thoracic CT (Calcification) |  |  |  |
| Thoracic Agatston Score, HU | 3333 (1207, 8622) | 1956 (369, 5486) | 0.26 |
| Calcification volume Score, HU | 4558 (1093, 8622) | 2543 (550, 7174) | 0.21 |
| -Central Blood pressure Index |  |  |  |
| Reflection Magnitude, % | 65 (59, 70) | 69 (61, 75) | 0.025 |
| Augmentation press., mmHg | 7.0 (4.0, 12.5) | 8.0 (5.0, 17.0) | 0.032 |
| Augmentation index, % | 17.0 (9.0, 30.0) | 24.5 (17.0, 38.0) | 0.001 |
| PWV, m/s | 10.5 (9.5, 12.5) | 11.2 (9.9, 12.6) | 0.12 |
| Central SBP, mmHg | 112.7 ± 17.4 | 112.7 ± 17.0 | 0.47 |
| Central DBP, mmHg | 78.9 ± 12.8 | 74.0 ± 13.0 | 0.018 |
| Central Pulse Pressure, mmHg | 55.2 ± 15.5 | 58.4 ± 15.2 | 0.19 |
| -Cardiac Echo |  |  |  |
| LVDd, mm | 45.0 ± 4.2 | 40.3 ± 5.2 | <0.0001 |
| LVDs, mm | 29.2 ± 4.5 | 25.0 ± 4.3 | <0.0001 |
| IVST, mm | 10.3 ± 1.5 | 9.8 ± 1.6 | 0.023 |
| PWT, mm | 10.2 ± 1.6 | 9.5 ± 1.5 | 0.006 |
| LVMI, g/m<sup>2</sup> | 93.5 (77.9, 107.0) | 80.5 (71.3, 100.0) | 0.028 |
| Relative Wall Thickness | 0.42 (0.40, 0.47) | 0.44 (0.40, 0.49) | 0.11 |
| Septal e' | 6.5 (5.8, 9.0) | 6.0 (4.8, 7.3) | 0.006 |
| Septal E/e' ratio | 10.8 (8.0, 15.2) | 12.4 (9.6, 15.5) | 0.029 |
| TR velocity | 2.5 (2.3, 2.9) | 2.6 (2.3, 2.8) | <0.0001 |
| LVEF (teichholz), % | 65.9 (59.3, 71.5) | 69.1 (64.0, 74.7) | 0.53 |
| E/A ratio, | 0.83 ± 0.34 | 0.75 ± 0.23 | 0.14 |
| Dct time, mmsec | 215.4 ± 66.8 | 232.1 ± 70.1 | 0.13 |
| LA dimension, mm | 37.5 ± 8.0 | 35.6 ± 7.9 | 0.14 |
| -Smoking habit |  |  |  |
| Habitual smoking, n (%) | 41 (60.3 %) | 8 (8.9 %) | <0.0001 |
| -Complication |  |  |  |
| HFpEF, n (%) | 16 (23.5 %) | 36 (40.0 %) | 0.029 |
| Hypertension, n (%) | 38 (55.9 %) | 62 (68.9 %) | 0.09 |
| Diabetes mellitus, n (%) | 23 (33.8 %) | 16 (17.8 %) | 0.021 |
| Hyperlipidemia, n (%) | 11 (16.2%) | 25 (27.8 %) | 0.09 |
| Af history, n (%) | 22 (32.4 %) | 27 (30.0 %) | 0.75 |
| Osteoporosis, n (%) | 7/24 (29.2%) | 33/52 (63.5%) | 0.005 |
| Hip total BMD, 0.75(YAM80%) | 87.2 ± 20.1 | 68.2 ± 17.1 | <0.0001 |

Abbreviations are the same as shown in Table 1.

Diabetes rate was significantly higher in males than in females (23/68 (33.8%) vs 16/90 (17.8%), p=0.021). DM patients presented significantly increased aortic calcification scores compared with non-DM patients: Agatston score (3889 (1911, 8031) vs (1412 (369, 5771) HU, p=0.040) and aortic calcium volume score (5883 (2795, 10729) vs (2047 (550, 7195) HU, p=0.021), although there was no significant difference in PWV (11.2 (10.2, 12.5) vs 10.9 (9.4, 12.7), p=0.31, respectively) in this study.

### Regression Analyses

The results of single regression analyses for HFpEF are shown in Table 3. The following factors: age; PWV; central systolic BP; Agatston Score; FGF23 plasma level; and, hypertension all revealed significant positive relations with HFpEF. Plasma levels of VD3 as well as of eGFR and male sex had significant negative relations with HFpEF. Phosphate serum level had significant regressions with male sex and plasma level of FGF23 (t=-2.44, p=0.016 and t=3.32, p=0.001, respectively).

**Table 3.**
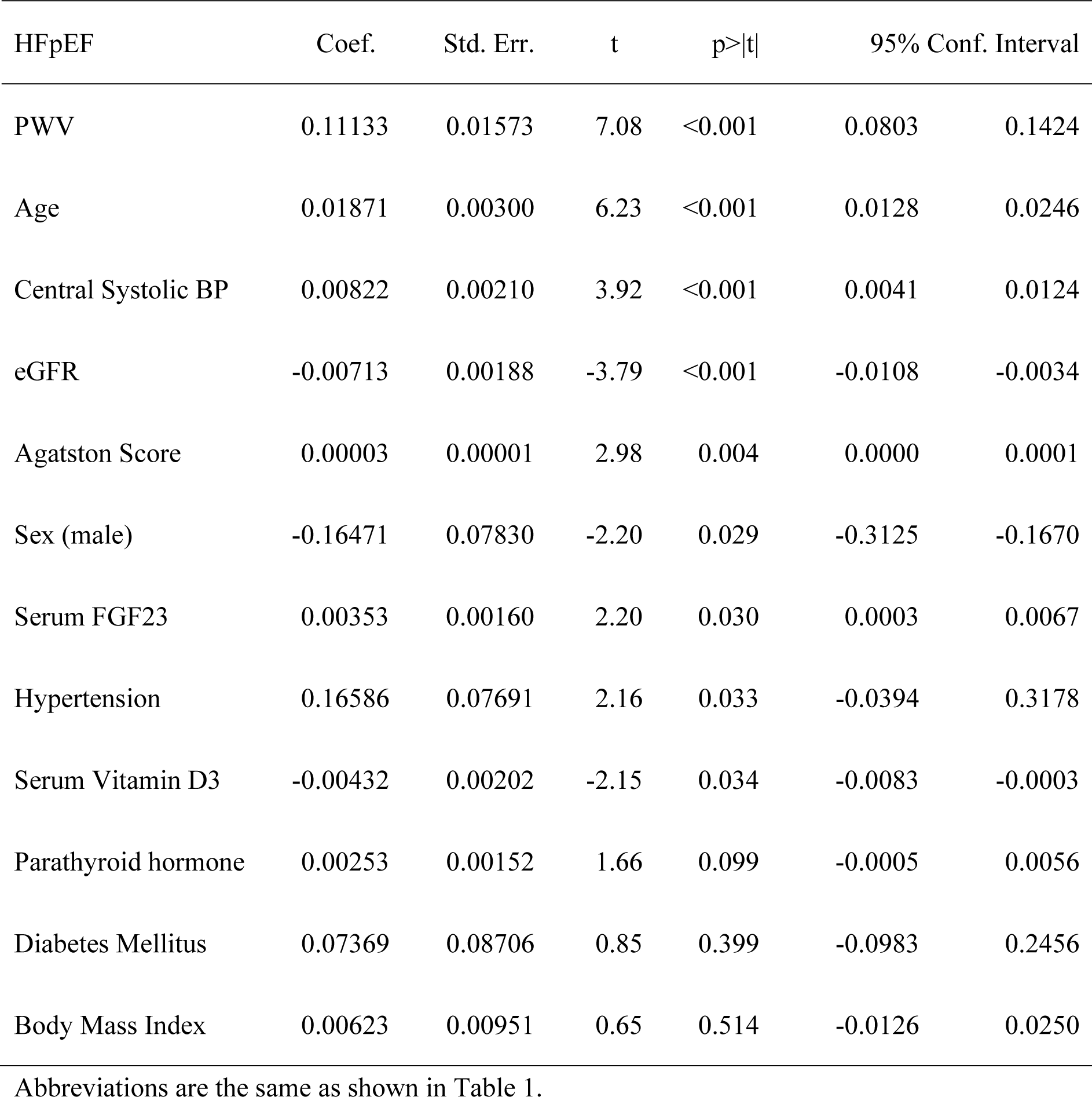
Single linear regression analyses for HFpEF.

| HFpEF | Coef. | Std. Err. | t | p>\|t\| | 95% Conf. Interval |  |
| --- | --- | --- | --- | --- | --- | --- |
| PWV | 0.11133 | 0.01573 | 7.08 | <0.001 | 0.0803 | 0.1424 |
| Age | 0.01871 | 0.00300 | 6.23 | <0.001 | 0.0128 | 0.0246 |
| Central Systolic BP | 0.00822 | 0.00210 | 3.92 | <0.001 | 0.0041 | 0.0124 |
| eGFR | -0.00713 | 0.00188 | -3.79 | <0.001 | -0.0108 | -0.0034 |
| Agatston Score | 0.00003 | 0.00001 | 2.98 | 0.004 | 0.0000 | 0.0001 |
| Sex (male) | -0.16471 | 0.07830 | -2.20 | 0.029 | -0.3125 | -0.1670 |
| Serum FGF23 | 0.00353 | 0.00160 | 2.20 | 0.030 | 0.0003 | 0.0067 |
| Hypertension | 0.16586 | 0.07691 | 2.16 | 0.033 | -0.0394 | 0.3178 |
| Serum Vitamin D3 | -0.00432 | 0.00202 | -2.15 | 0.034 | -0.0083 | -0.0003 |
| Parathyroid hormone | 0.00253 | 0.00152 | 1.66 | 0.099 | -0.0005 | 0.0056 |
| Diabetes Mellitus | 0.07369 | 0.08706 | 0.85 | 0.399 | -0.0983 | 0.2456 |
| Body Mass Index | 0.00623 | 0.00951 | 0.65 | 0.514 | -0.0126 | 0.0250 |

Abbreviations are the same as shown in Table 1.

Multivariable logistic regression analysis finally was done for dependent variable, HFpEF. There were significant collinearities as follows: between age and PWV (t=33.62, p<0.001); between PWV and eGFR (t=-6.34, p<0.001); between PWV and FGF23 (t=3.33, p<0.001); between PWV and vitamin D3 (t=-2.29, p=0.024); and, between FGF23 and vitamin D3 (t=-4.82, p<0.001), respectively. PWV higher values, hypertension and male sex were selected as independent variables. The analysis revealed that PWV higher values (t=5.89, p<0.0001) and hypertension (t=2.17, p=0.031) were significant predictors for HFpEF. (Table 4)

**Table 4.**
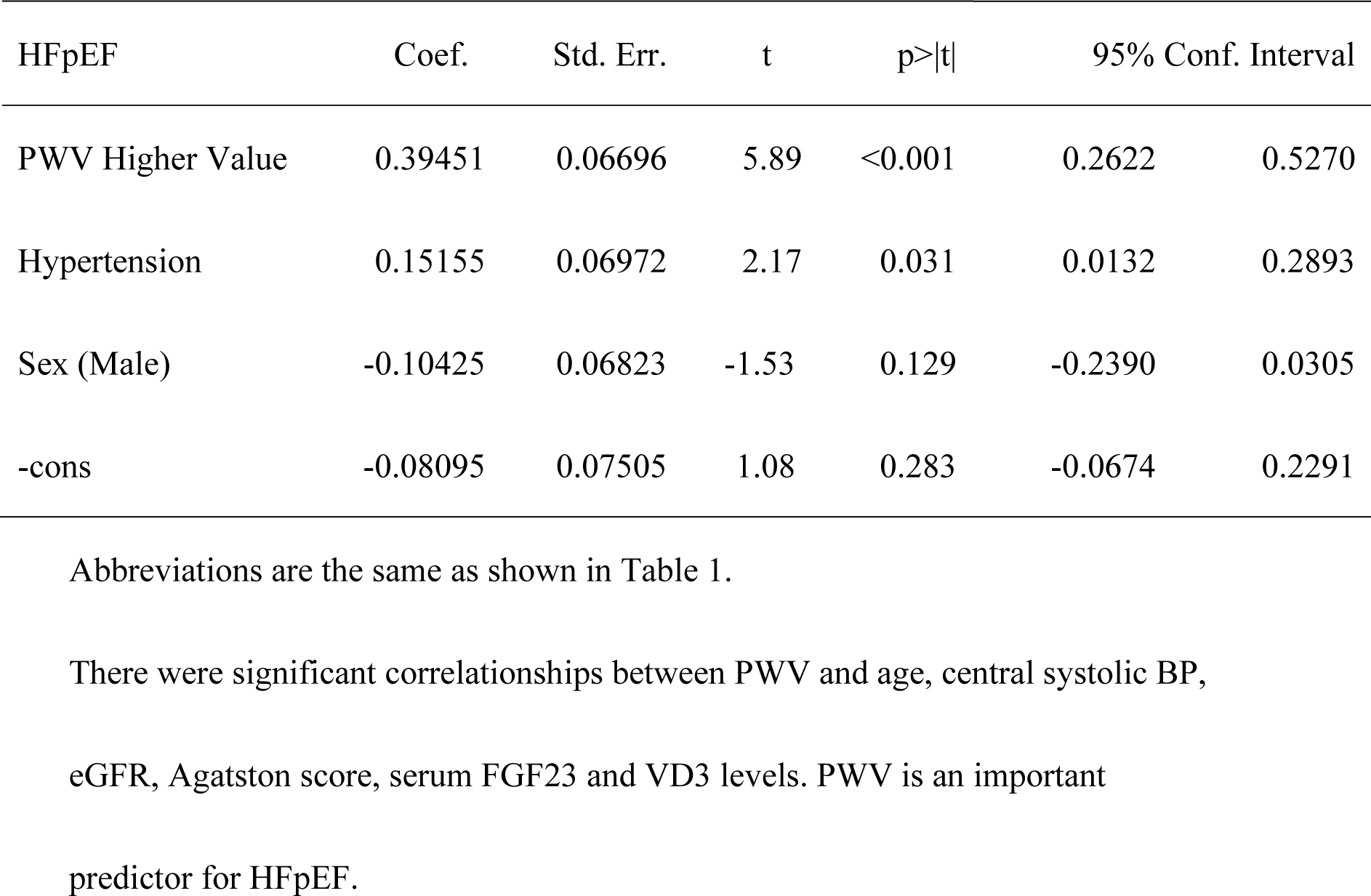
Multiple logistic linear regression analyses for HFpEF n=158, F=(3, 154)=15.03, p<0.0001, R-squared=0.2265.

| HFpEF | Coef. | Std. Err. | t | p>\|t\| | 95% Conf. Interval |  |
| --- | --- | --- | --- | --- | --- | --- |
| PWV Higher Value | 0.39451 | 0.06696 | 5.89 | <0.001 | 0.2622 | 0.5270 |
| Hypertension | 0.15155 | 0.06972 | 2.17 | 0.031 | 0.0132 | 0.2893 |
| Sex (Male) | -0.10425 | 0.06823 | -1.53 | 0.129 | -0.2390 | 0.0305 |
| -cons | -0.08095 | 0.07505 | 1.08 | 0.283 | -0.0674 | 0.2291 |

Abbreviations are the same as shown in Table 1.
There were significant correlations between PWV and age, central systolic BP, eGFR, Agatston score, serum FGF23 and VD3 levels. PWV is an important predictor for HFpEF.

## DISCUSSION

Heart failure with preserved ejection fraction (HFpEF) is increasingly recognized as a major public health concern worldwide.^1-4^ Some HFpEF patients present with obesity and/or diabetes mellitus (DM) which often results in atherosclerosis and LV hypertrophy.^1,2,5,6^ Another phenotype of HFpEF is frequently seen in elderly patients with CKD, as Cohen et al. reported.^1,2^ Elderly people with CKD frequently present with aortic calcification similar to hemodialysis patients.^9-11^ We recently reported that a deterioration of phosphate homeostasis (and/or phosphate retention) obtained by measuring the serum levels of FGF23 and VD3, leads to arterial calcification, and could produce arterial stiffness which intensifies cardiac afterload.^26^ We therefore investigated whether arterial stiffness due to phosphate retention could lead to HFpEF.

Table 1 revealed that the HFpEF group indicated the following characteristics: higher age; female sex; renal dysfunction; higher systolic BP; higher pulse pressure; inflammation; higher BNP levels; higher aortic calcification scores; higher PWV; higher central pulse pressure; and, higher cardiac afterload markers, including augmentation pressure and central systolic BP. The HFpEF group further presented hypertension associated factors and diastolic dysfunction markers, as documented in the 2021 ESC guidelines: LV thickness; concentric hypertrophy; decreased e’; increased E/e’; LA larger dimension; and a history of atrial fibrillation.^4^ The HFpEF group moreover had increasing plasma levels of FGF23 and decreasing plasma levels of VD3. Here, we can therefore see that the phosphate retention effects on arterial stiffness^25^ and the pathological mechanism of HFpEF are related.

Vlachopoulos et al. reported that PWV is a gold standard for measuring arterial stiffness, and that arterial stiffness is causative of pulsatile afterload.^14,28^ Arterial wave reflections increase according to the degree of arterial stiffness, leading to the incrementation of mid-to late systolic load and subsequent LV abnormalities, including LV concentric remodeling and myocardial hypertrophy.^14,28^

Arterial stiffness was intensified according to age and renal dysfunction degree, as is shown in Figure 1. The plasma levels of FGF23 and VD3 dovetailed with age and renal dysfunction as we and others have presented.^17,20-26^ The loss of nephrons due to aging and CKD causes a phosphate excretion disorder in the renal proximal tubules.^10,17-23^

Phosphate retention is an important cause of arterial calcification.^17-23^ Once the concentration of calcium and phosphate ions exceeds the blood saturation level, because the extracellular fluid is super-saturated in terms of phosphate and calcium ions, therefore an increase in the phosphate concentration can trigger precipitation of calcium-phosphate.^17,20,37,38^ Calcium-phosphate precipitated upon an increase in the blood phosphate concentration and is then absorbed by serum protein fetuin-A to form colloidal nanoparticles called calciprotein particles (CPPs). CPPs in the blood can induce cell damage, ectopic calcification, and inflammatory responses.^17,20,37,38^

Increased FGF23 increases phosphate excretion per nephron, it therefore compensates for any reduced nephron number and as a result it maintains phosphate homeostasis.^20^ Increased phosphaturia is independently associated with a decline of eGFR in stage 2-3 CKD patients with normal blood phosphate levels.^39^ Phosphate retention, which is a trigger for arterial calcification, may be launched in the earliest stages of the aging process.

This study, further presents that the more arterial stiffness there is, the more augmentation pressure increases and the more central systolic BP increases, both of which are markers of cardiac afterload. (Figure 2) This intensified cardiac afterload, indicated by central systolic BP, had significant relationships with LV hypertrophy, which leads to LV diastolic dysfunction (e’ and E/e’) as we presented in our results section. Leite et al. reported that LV diastolic dysfunction is induced by an increased afterload in the healthy hearts of rabbits and dogs.^40^ Roy et al. reported that plasma level of FGF23 was increased in patients with HFpEF and it was associated with a low survival rate.^41^ By indicating the relationship between PWV and HFpEF judgement parameters, our study suggests that arterial stiffness leads to LV concentric hypertrophy, and finally to HFpEF. (Figures 3 and 4).

Table 2 shows that elderly females, rather than males, more commonly present with HFpEF.^1,2,9^ Elderly females demonstrated significant characteristics of arterial stiffness shown by an increased percent of AI, compared with males, as Goto et al. and we presented.^9^ Females showed significantly higher levels of serum phosphate, and lower bone mineral density levels than those in males. Bone is a safe and important depository for phosphate.^10,15-18^ Osteoporosis patients have disadvantages in the management of phosphate; therefore, elderly females may have more arterial stiffness and HFpEF than males.

The diabetes mellitus rate is not different between the HFpEF and the non-HFpEF groups, although there were relatively few patients with DM and/or obesity. DM patients had higher Agatston and calcification volume scores than non-DM patients in this study. Cohen et al. showed that obese and diabetic patients had a higher pulsatile arterial load, increased concentric LV hypertrophy and an increasing of E/e’.^2,6^ Chirinos et al. reported in HFpEF patients with DM there was an increasing of arterial stiffness, PWV and LV mass.^6^ DM or obesity, as well as elderly people with CKD, all have increased PWV values.^6,9,14^ It is natural to assume therefore that arterial stiffness should be considered an important mechanism of HFpEF.^6,9-14,26^

Single linear regression analyses showed that PWV, age, central systolic BP, eGFR, Agatston calcification scores, female, serum FGF23 levels, hypertension, and serum VD3 levels, all revealed significant relationships for the dependent factor, HFpEF. All these factors are related to phosphate retention or arterial stiffness. There were many significant collinearities in the phosphate retention related factors. These mean that phosphate retention is strongly related to HFpEF. We determined PWV, hypertension and sex difference were independent factors for HFpEF by considering their collinearity. Multivariable logistic regression analyses revealed that both PWV and hypertension were important predictors for HFpEF. This study reveals that the main triggering mechanism of HFpEF is arterial stiffness effected by phosphate retention.

The mechanism and/or causation of HFpEF is clearly different from that of heart failure with reduced ejection fraction (HFrEF) which is a result of myocardial dysfunction. This study presents that phosphate retention, due to a loss of nephrons associated with aging and CKD, accelerates the aging process. Increasing arterial stiffness leads to LV diastolic dysfunction and/or HFpEF by amplifying cardiac afterload.

It is important to recognize phosphate retention using FGF23 and VD3 measurements in the early stages of CKD. An increase in the former and a decrease in the latter indicate the actual degree of phosphate retention. We propose that Klotho gene up-regulation therapy, phosphate regulating medications, phosphate restriction diets, and osteoporosis therapies may all be effective for phosphate control, especially in females.^42,43^ Preventing of phosphate retention should be a new clinical target for treating aging and aging related diseases.^17,20,21,27,42,43^

### Limitations

This is a cross-sectional observation study. There is a limited discussion of causes and effects. Our patients were mainly elderly patients suspected of heart failure.

There were no patients who needed invasive hemodynamic measurements during exercise. There was only a limited discussion about younger patients and obese or diabetic patients. We used BaPWV tests to estimate central blood pressure, especially afterload related markers. Recently cardio-ankle vascular index has been used by some as a marker for arterial stiffness.

## Conclusions

Arterial stiffness degree increases due to aging and CKD; furthermore, it is an important trigger for HFpEF. Arterial stiffness produces cardiac afterload which leads to LV concentric hypertrophy and LV diastolic dysfunction. PWV, as well as hypertension, is an important predictor for HFpEF. Arterial stiffness, due to phosphate retention, should be a new therapeutic target for fighting aging related diseases, as well as preventing cardiac mortality and morbidity, including HFpEF.

## Data Availability

No problem

## REFERENCES

1. Lam CS, Donal E, Kraigher-Krainer E, Vasan RS. Epidemiology and clinical course of heart failure with preserved ejection fraction. Eur J Heart Fail. 2011;13:18–28.

2. Cohen JB, Schrauben SJ, Zhao L, Basso MD, Cvijic ME, Li Z, Yarde M, Wang Z, Bhattacharya PT, Chirinos DA, et al. Clinical phenogroups in heart failure with preserved ejection fraction: detailed phenotypes, prognosis, and response to spironolactone. JACC Heart Fail. 2020;8:172–184.

3. Heidenreich PA, Bozkurt B, Aguilar D, Allen LA, Byun JJ, Colvin MM, Deswal A, Drazner MH, Dunlay SM, Evers LR, et al. 2022 AHA/ACC/HFSA guideline for the management of heart failure: a report of the American College of Cardiology/American Heart Association joint committee on clinical practice guidelines. Circulation. 2022;145:e895–e1032.

4. McDonagh TA, Metra M, Adamo M, Gardner RS, Baumbach A, Böhm M, Burri H, Butler J, Čelutkienė J, Chioncel O, et al. 2021 ESC guidelines for the diagnosis and treatment of acute and chronic heart failure: developed by the task force for the diagnosis and treatment of acute and chronic heart failure of the European Society of Cardiology (ESC). with the special contribution of the heart failure association (HFA) of the ESC. Eur J Heart Fail. 2022;24:4–131.

5. Meagher P, Adam M, Civitarese R, Bugyei-Twum A, Connelly KA. Heart failure with preserved ejection fraction in diabetes: mechanisms and management. Can J Cardiol. 2018;34:632–643.

6. Chirinos JA, Bhattacharya P, Kumar A, Proto E, Konda P, Segers P, Akers SR, Townsend RR, Zamani P. Impact of diabetes mellitus on ventricular structure, arterial stiffness, and pulsatile hemodynamics in heart failure with preserved ejection fraction. J Am Heart Assoc. 2019;8:e011457.

7. Maslov PZ, Kim JK, Argulian E, Ahmadi A, Narula N, Singh M, Bax J, Narula J. Is cardiac diastolic dysfunction a part of post-menopausal syndrome? JACC Heart Fail. 2019;7:192–203.

8. Kasiakogias A, Rosei EA, Camafort M, Ehret G, Faconti L, Ferreira JP, Brguljan J, Januszewicz A, Kahan T, Manolis A, et al. Hypertension and heart failure with preserved ejection fraction: position paper by the European Society of Hypertension. J Hypertens. 2021;39:1522–1545.

9. Goto T, Ohte N, Fukuta H, Wakami K, Tani T, Kimura G. Relationship between effective arterial elastance, total vascular resistance, and augmentation index at the ascending aorta and left ventricular diastolic function in older women. Circ J. 2013;77:123–129.

10. Nelson AJ, Raggi P, Wolf M, Gold AM, Chertow GM, Roe MT. Targeting vascular calcification in chronic kidney disease. JACC Basic Transl Sci. 2020;5:398–412.

11. Fujiu A, Ogawa T, Matsuda N, Ando Y, Nitta K. Aortic arch calcification and arterial stiffness are independent factors for diastolic left ventricular dysfunction in chronic hemodialysis patients. Circ J. 2008;72:1768–1772.

12. Mottram PM, Haluska BA, Leano R, Carlier S, Case C, Marwick TH. Relation of arterial stiffness to diastolic dysfunction in hypertensive heart disease. Heart. 2005;91:1551–1556.

13. Ooi H, Chung W, Biolo A. Arterial stiffness and vascular load in heart failure. Congest Heart Fail. 2008;14:31–36.

14. Weber T, O’Rourke MF, Ammer M, Kvas E, Punzengruber C, Eber B. Arterial stiffness and arterial wave reflections are associated with systolic and diastolic function in patients with normal ejection fraction. Am J Hypertens. 2008;21:1194–1202.

15. Stenvinkel P, Larsson TE. Chronic kidney disease: a clinical model of premature aging. Am J Kidney Dis. 2013;62:339–351.

16. Kuro-o M, Matsumura Y, Aizawa H, Kawaguchi H, Suga T, Utsugi T, Ohyama Y, Kurabayashi M, Kaname T, Kume E, et al. Mutation of the mouse klotho gene leads to a syndrome resembling ageing. Nature. 1997;390:45–51.

17. Kuro OM. Phosphate as a Pathogen of Arteriosclerosis and Aging. J Atheroscler Thromb. 2021;28:203–213.

18. Gross P, Six I, Kamel S, Massy ZA. Vascular toxicity of phosphate in chronic kidney disease: beyond vascular calcification. Circ J. 2014;78:2339–2346.

19. Ärnlöv J, Carlsson AC, Sundström J, Ingelsson E, Larsson A, Lind L, Larsson TE. Higher fibroblast growth factor-23 increases the risk of all-cause and cardiovascular mortality in the community. Kidney Int. 2013;83:160–166.

20. Kuro OM. Aging and FGF23-klotho system. Vitam Horm. 2021;115:317–332.

21. Haussler MR, Whitfield GK, Kaneko I, Forster R, Saini R, Hsieh JC, Haussler CA, Jurutka PW. The role of vitamin D in the FGF23, klotho, and phosphate bone-kidney endocrine axis. Rev Endocr Metab Disord. 2012;13:57–69.

22. Bergwitz C, Jüppner H. Regulation of phosphate homeostasis by PTH, vitamin D, and FGF23. Annu Rev Med. 2010;61:91–104.

23. Jacquillet G, Unwin RJ. Physiological regulation of phosphate by vitamin D, parathyroid hormone (PTH) and phosphate (Pi). Pflugers Arch. 2019;471:83–98.

24. Kassi E, Adamopoulos C, Basdra EK, Papavassiliou AG. Role of vitamin D in atherosclerosis. Circulation. 2013;128:2517–2531.

25. Wang TJ, Pencina MJ, Booth SL, Jacques PF, Ingelsson E, Lanier K, Benjamin EJ, D’Agostino RB, Wolf M, Vasan RS. Vitamin D deficiency and risk of cardiovascular disease. Circulation. 2008;117:503–511.

26. Mizuno Y, Ishida T, Kugimiya F, Takai S, Nakayama Y, Yonemitsu K, Harada E. Deterioration of phosphate homeostasis is a trigger for cardiac afterload - clinical importance of fibroblast growth factor 23 for accelerated aging. Circ Rep. 2023;5:4–12.

27. Lang RM, Badano LP, Mor-Avi V, Afilalo J, Armstrong A, Ernande L, Flachskampf FA, Foster E, Goldstein SA, Kuznetsova T, et al. Recommendations for cardiac chamber quantification by echocardiography in adults: an update from the American Society of Echocardiography and the European Association of Cardiovascular Imaging. Eur Heart J Cardiovasc Imaging. 2015;16:233–270.

28. Vlachopoulos C, Hirata K, O’Rourke MF. Pressure-altering agents affect central aortic pressures more than is apparent from upper limb measurements in hypertensive patients: the role of arterial wave reflections. Hypertension. 2001;38:1456–1460.

29. Wei W, Tölle M, Zidek W, van der Giet M. Validation of the mobil-O-Graph: 24 h-blood pressure measurement device. Blood Press Monit. 2010;15:225-228.

30. Paiva AMG, Mota-Gomes MA, Brandão AA, Silveira FS, Silveira MS, Okawa RTP, Feitosa ADM, Sposito AC, Nadruz W, Jr. Reference values of office central blood pressure, pulse wave velocity, and augmentation index recorded by means of the Mobil-O-Graph PWA monitor. Hypertens Res. 2020;43:1239–1248.

31. Craiem D, Casciaro M, Pascaner A, Soulat G, Guilenea F, Sirieix ME, Simon A, Mousseaux E. Association of calcium density in the thoracic aorta with risk factors and clinical events. Eur Radiol. 2020;30:3960–3967.

32. Yamazaki Y, Okazaki R, Shibata M, Hasegawa Y, Satoh K, Tajima T, Takeuchi Y, Fujita T, Nakahara K, Yamashita T et al. Increased circulatory level of biologically active full-length FGF-23 in patients with hypophosphatemic rickets/osteomalacia. J Clin Endocrinol Metab. 2002;87:4957–4960.

33. Fraser WD, Durham BH, Berry JL, Mawer EB. Measurement of plasma 1,25 dihydroxyvitamin D using a novel immunoextraction technique and immunoassay with iodine labelled vitamin D tracer. Ann Clin Biochem. 1997;34 (Pt 6):632–637.

34. Gao P, Scheibel S, D’Amour P, John MR, Rao SD, Schmidt-Gayk H, Cantor TL. Development of a novel immunoradiometric assay exclusively for biologically active whole parathyroid hormone 1-84: implications for improvement of accurate assessment of parathyroid function. J Bone Miner Res. 2001;16:605–614.

35. Yasue H, Yoshimura M, Sumida H, Kikuta K, Kugiyama K, Jougasaki M, Ogawa H, Okumura K, Mukoyama M, Nakao K. Localization and mechanism of secretion of B-type natriuretic peptide in comparison with those of A-type natriuretic peptide in normal subjects and patients with heart failure. Circulation. 1994;90:195–203.

36. Katz MH. Multivariable Analysis: A Practical Guide for Clinicians and Public Health Researchers. 3rd ed. Cambridge: Cambridge University Press; 2011.

37. Akiyama KI, Miura Y, Hayashi H, Sakata A, Matsumura Y, Kojima M, Tsuchiya K, Nitta K, Shiizaki K, Kurosu H, et al. Calciprotein particles regulate fibroblast growth factor-23 expression in osteoblasts. Kidney Int. 2020;97:702–712.

38. Kunishige R, Mizoguchi M, Tsubouchi A, Hanaoka K, Miura Y, Kurosu H, Urano Y, Kuro OM, Murata M. Calciprotein particle-induced cytotoxicity via lysosomal dysfunction and altered cholesterol distribution in renal epithelial HK-2 cells. Sci Rep. 2020;10:20125.

39. Santamaría R, Díaz-Tocados JM, Pendón-Ruiz de Mier MV, Robles A, Salmerón-Rodríguez MD, Ruiz E, Vergara N, Aguilera-Tejero E, Raya A, Ortega R, et al. Increased phosphaturia accelerates the decline in renal function: a search for mechanisms. Sci Rep. 2018;8:13701.

40. Leite S, Rodrigues S, Tavares-Silva M, Oliveira-Pinto J, Alaa M, Abdellatif M, Fontoura D, Falcão-Pires I, Gillebert TC, Leite-Moreira AF, Lourenço AP. Afterload-induced diastolic dysfunction contributes to high filling pressures in experimental heart failure with preserved ejection fraction. Am J Physiol Heart Circ Physiol. 2015;309:H1648–654.

41. Roy C, Lejeune S, Slimani A, de Meester C, Ahn As SA, Rousseau MF, Mihaela A, Ginion A, Ferracin B, Pasquet A, et al. Fibroblast growth factor 23: a biomarker of fibrosis and prognosis in heart failure with preserved ejection fraction. ESC Heart Fail. 2020;7:2494–2507.

42. Wang J, Zhou JJ, Robertson GR, Lee VW. Vitamin D in vascular calcification: a double-edged sword? Nutrients. 2018;10.

43. Shigematsu T, Sonou T, Ohya M, Yokoyama K, Yoshida H, Yokoo T, Okuda K, Masumoto AR, Iwashita Y, Iseki K, et al. Preventive strategies for vascular calcification in patients with chronic kidney disease. Contrib Nephrol. 2017;189:169–177.

